# Sustainable integration of a vertical voluntary medical male circumcision program into routine health services in Zimbabwe: A mixed methods evaluation of a participatory change intervention

**DOI:** 10.1101/2024.09.06.24313083

**Authors:** Amanda Marr Chung, Joseph Murungu, Precious Chitapi, Rudo Chikodzore, Peter Case, Jonathan Gosling, Roly Gosling, Sinokuthemba Xaba, Getrude Ncube, Owen Mugurungi, Patience Kunaka, Stefano M. Bertozzi, Caryl Feldacker

## Abstract

The global health community has recognized the importance of integrating and sustaining health programs and forming equitable partnerships. Corresponding with these objectives, international aid donors are embracing the principle of localization. The Voluntary Medical Male Circumcision (VMMC) in Zimbabwe is a large vertical HIV prevention program primarily funded through development assistance for health. Program stakeholders want to sustainably integrate VMMC into routine health services so that the program will continue to be a cost-effective HIV prevention strategy through 2030. The purpose of this paper is to describe a bottom-up process of sustainably integrating the program into routine health services through an approach that empowers local stakeholders. At the district level, we facilitated changes to accelerate integration and sustainability. To evaluate our intervention, we used a mixed methods design comprising analysis of district-level work plans with qualitative and quantitative indicators, combined with a survey assessing sustainability capacity of the program, administered at midline and endline to district teams. In all five pilot districts we facilitated the transition of VMMC into the government’s district administration, resulting in a locally owned and managed program, while also strengthening individual and team capacity. We observed improvements across all World Health Organization health system building blocks, suggesting that the intervention strengthened the overall health system. The sustainability survey showed a reduction in funding stability but a significant increase in communications, program adaptation, and organizational capacity. Compared to traditional top-down change initiatives, the participatory approach to integration was an effective way of addressing specific VMMC challenges at the district level whilst maintaining management and oversight at provincial and national levels. Other health programs in low- and middle-income countries seeking to integrate and sustain health services at subnational levels should consider this diagonal, bottom-up model to promote local leadership development and health system strengthening.

## Introduction

Despite growing emphasis on sustainability and integration in health programs major funders such as the United States Government and the Global Fund to Fight AIDS, Tuberculosis, and Malaria continue to make large investments in vertical health programs in countries that rely on international development assistance for health (DAH) [1–6]. While we saw record spending for COVID-19 in 2020-2021, DAH for many other health areas did not grow [7]. Additionally, domestic governments of low- and middle-income countries face increasing pressure to allocate at least 15% of their government spending towards health expenditure [7]. International aid donors are also embracing localization, i.e., the shifting of power and funding to local partners, a strategy aligned with the broader goals of integrating and sustaining health programs and encouraging equitable partnerships in global health.

Voluntary Medical Male Circumcision (VMMC) in Zimbabwe is delivered through a national HIV prevention program, largely funded through DAH. According to 2022 UNAIDS estimates, Zimbabwe has 1.3 million adults and children living with HIV [8]. The World Health Organization (WHO) and UNAIDS designated Zimbabwe as a priority country for voluntary male medical circumcision (VMMC) due to the country having one of the highest adult HIV prevalence in the world (11%), with unprotected heterosexual sex as a driver of HIV transmission [9]. The VMMC program is transitioning from a vertical to a horizontal program. A vertical health program focuses on addressing a single disease and is funded and managed by external donors and partners, operating independently from the routine health system. In contrast, a horizontal health program is publicly financed, integrated into the health system, and aims to improve overall health outcomes. As of 2024, the VMMC program is in a hybrid state of transition: although all 63 districts are government run, most are funded by external donors and supported by implementing partners (IPs). The transition to sustainability was guided by a Sustainability Transition Implementation Plan (STIP) that prioritizes integration, decentralization, and local ownership of the program. According to the STIP, a fully sustainable VMMC program will have reached an ideal stage when it achieves:

> The managerial, financial, and operational ability to deliver and maintain 80% voluntary medical male circumcision coverage to ensure long-term health benefits and reduction in new HIV infections. This is achieved through conformity to social norms, local ownership rendering the programme affordable, accessible and acceptable to all. [10]

Since 2020, a shift in policy to prioritize the 15-29 years age group, due to concerns around the number of adverse events in young boys, resulted in a reduction of the pool of males available to be circumcised. From 2013-2017, boys aged 10 to 14 years contributed almost 50% of all male circumcisions [10].

### Progress towards integration and sustainability

Before 2020, certain IPs had already initiated the integration of the VMMC program into routine health services. From program inception, the PEPFAR-funded ZAZIC Consortium took steps toward integrating the VMMC program into MoHCC facilities, collaborating with MoHCC teams across 21 districts starting in 2013 [11]. Vu et al. provided a comprehensive account of their recommendations and insights gained during the transition from a USA-based organization to local management and ownership of these PEPFAR-funded districts [12]. However, in the majority of the remaining districts, the process of transitioning to sustainability did not commence until 2018. To monitor the national transition to sustainability, in 2019 the MoHCC worked with Clinton Health Access Initiative (CHAI) to develop a VMMC Transition Assessment Dashboard (VTAD).

### Current state of VMMC programs

Beginning in 2008, VMMC was rolled out as a fully donor-funded operation, forming part of the emergency response to HIV in fifteen eastern and southern African countries [13]. Over the past fifteen years, these VMMC programs successfully reduced HIV risk in male and female populations, with projections to avert at least 4.5 million new HIV infections by 2050 [13]. Moreover, VMMC provides additional health benefits to both males and females, reducing the risk of STIs such as HPV, bacterial vaginosis, herpes simplex virus-2, and Trichomonas vaginalis, as well as lowering the risks of cervical, prostate, and penile cancers [14–16].

In the current stage of the HIV epidemic, VMMC programs are pressed to change in response to three major factors:

1) The target population for VMMC programs now comprises adolescents/young men (ages 15-29 years), particularly in areas where older sexually active males have already been circumcised [13,17]. This shift in the target priority age group implies a reduction in the volume of VMMC procedures required, with fewer adolescents eligible for the procedure.
2) Major bilateral donors are reducing their contributions to AIDS financing [18]. While these donors have committed to continued investment in VMMC for the next five years, VMMC programs, along with other HIV prevention methods, face the challenge of maintaining adequate financing beyond this period [19].
3) Countries with VMMC programs aim to assume greater ownership of the program while integrating VMMC into their general health services, in accordance with the Paris Declaration of Aid Effectiveness of 2005 and the Accra Agenda for Action of 2008 [20]. The trend towards localization aligns with transition strategies of international aid, including the U.S. President’s Emergency Plan for AIDS Relief (PEPFAR) [21].

As countries strive to achieve universal health coverage, the lack of consensus surrounding the definitions of integration and sustainability hinders the progress of donor-funded global health programs towards this end. Both of these are essential components of bilateral donors’ strategies, such as PEPFAR’s new approach, and feature prominently in the policies of multilateral donors, including the Global Fund for AIDS, TB, and Malaria’s Sustainability, Transition, and Co-financing Policy [4,22].

Sustainability remains a donor-driven process. Due to the unstable economic situation, there are no immediate plans to fill any funding gaps left by donors with domestic funding. The World Health Organization (WHO) has identified a research gap concerning participatory, implementation approaches for integrating VMMC with other health services [17]. This paper aims to describe the processes and results of the OPTIMISE Project that used the Leadership and Engagement for Improved Accountability and Delivery of Services (LEAD) Framework to support sustainably integrating the vertical VMMC program into routine health services in Zimbabwe. We provide further details below of the LEAD Framework, a multi-faceted systems change approach combining organization development, quality improvement, and capacity strengthening. A key characteristic of the LEAD Framework lies in its application of participatory, bottom-up techniques that foster buy-in across the entire system, from community and district to provincial and national levels.

This research paper intends to contribute to the field in two major ways: (1) by offering empirical evidence of how VMMC services can be integrated into a health system in a participatory, sustainable manner; and (2) by introducing a bottom-up intervention methodology that can facilitate practical and effective integration, expediting the pathway to sustained VMMC implementation and support program integration in other health system settings.

## Methods

### Definitions

The terms ‘integration’ and ‘sustainability’ are closely interconnected. The ultimate long-term objective of an integrated program is to achieve local ownership and sustainability, ensuring that the program continues to function effectively even after external support is scaled back or ends [23]. However, sustainability is predicated on adequate funding to maintain the program’s operations and impact. Both integration and sustainability are also best considered as existing along a continuum rather than being viewed as binary outcomes [24]. A common feature of vertical programs is that they express the priorities of global health donors, which are not necessarily identical to the health priorities identified by local stakeholders. We define a vertical health program as one that focuses on a single disease or population group, while a horizontal program has a broader scope and longer term goals centered on primary care or general health service provision [25]. A diagonal approach tackles specific diseases through health system strengthening [26,27]. Finally, we employ the term ‘hybrid program’ to refer to the state of transition between a vertical and horizontal mode of service provision.

### The OPTIMISE project

The overarching goal of the two-year University of California, San Francisco (UCSF) OPTIMISE project was to enhance leadership and management capacity within the Ministry of Health and Child Care (MoHCC) to significantly change parts of the health system to improve integration and sustainability. An extension of this work is now being funded through a local Zimbabwean partner, Precious Innovations Ubuntu Global Charitable Trust. In 2020, the MoHCC sought support from UCSF’s LEAD team, complementing the efforts of CHAI and PSH, in transitioning the VMMC program to sustainability on national and subnational levels. The start of the project coincided with the COVID-19 pandemic, which significantly affected the majority of the 15 VMMC priority countries in eastern and southern Africa. As a result, these countries missed the cumulative 2016-2020 target of 25 million VMMCs by 7 million [28]. At its peak, Zimbabwe performed over 350,000 male circumcisions in 2019 [28].

### LEAD Framework

The study presented in this paper utilizes the LEAD Framework as the primary intervention method [29]. The LEAD team adapted the approach and tools for this project on those developed in a previous intervention for malaria elimination programs in Eswatini, Namibia, and Zimbabwe [30–32]. At the core of the LEAD Framework is the application of participatory action research (PAR) to improve the health system. It is based on the principle that the actors involved in delivery of outcomes are the ones best placed, with facilitative support, to identify problems and implement solutions and system changes [33].

Key components of PAR and the LEAD Framework include exercises to facilitate communication, teamwork, and problem identification and resolution through the iterative development of work plans and metrics to measure team progress (Fig 1). We strengthened organization development, leadership, and facilitation skills through a year-long university accredited postgraduate training program entitled ‘Professional Practice in Change Leadership’ (PPCL). We selected local facilitators for the training so that the work could be sustained when we withdrew our support. For details on the organizational development and quality improvement tools and techniques used to transition the VMMC program, refer to S1 Table.

**Fig 1.**
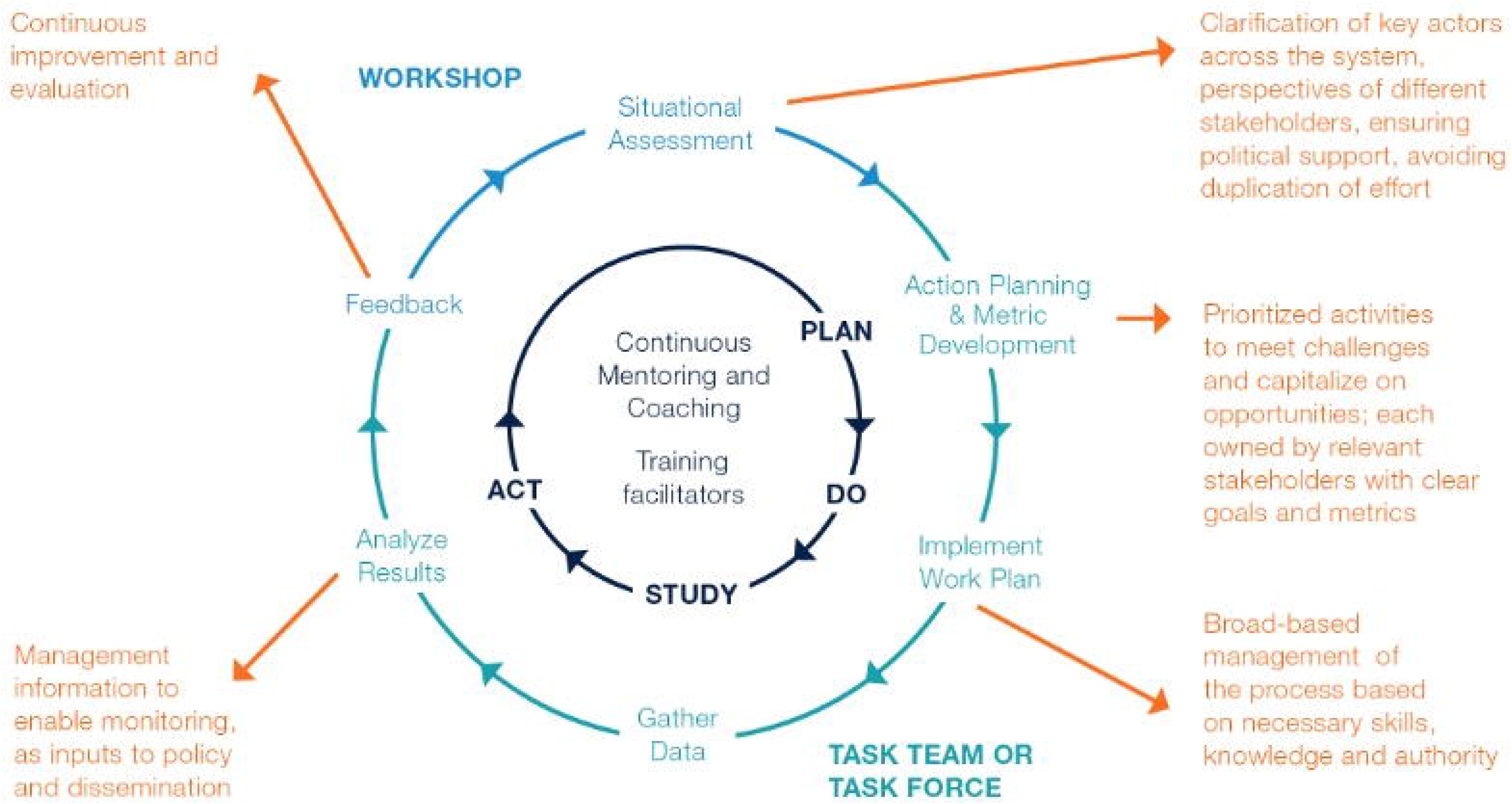
LEAD Framework cycle. The LEAD Framework cycle improves processes through the convening of stakeholders at all levels, action planning, and implementation through iterative Plan-Do-Study-Act cycles, continuous mentoring and coaching, training of facilitators, and feedback of results to support continuous improvement.

### Partnerships

The LEAD team that launched the OPTIMISE project included individuals from Zimbabwe, the United Kingdom, and the United States and was composed of a local Zimbabwean NGO, independent consultants, Women’s University in Africa, UCSF, and the University of West of England. The team included experts in global health, evaluation, facilitation, quality improvement, change management, leadership development and participatory action research. The LEAD team and the MoHCC collectively agreed on the goals of the OPTIMISE project, the overall approach to systems change, and engagement of the MoHCC at different levels of the health system. In implementing the OPTIMISE project, activities using LEAD took place over a two-year period through a national Task Force and at subnational levels through multisectoral and interdisciplinary district Task Teams. The cadence of meetings to develop work plans varied depending on the level of engagement. See Fig 2 for a timeline of project activities.

**Fig 2.**
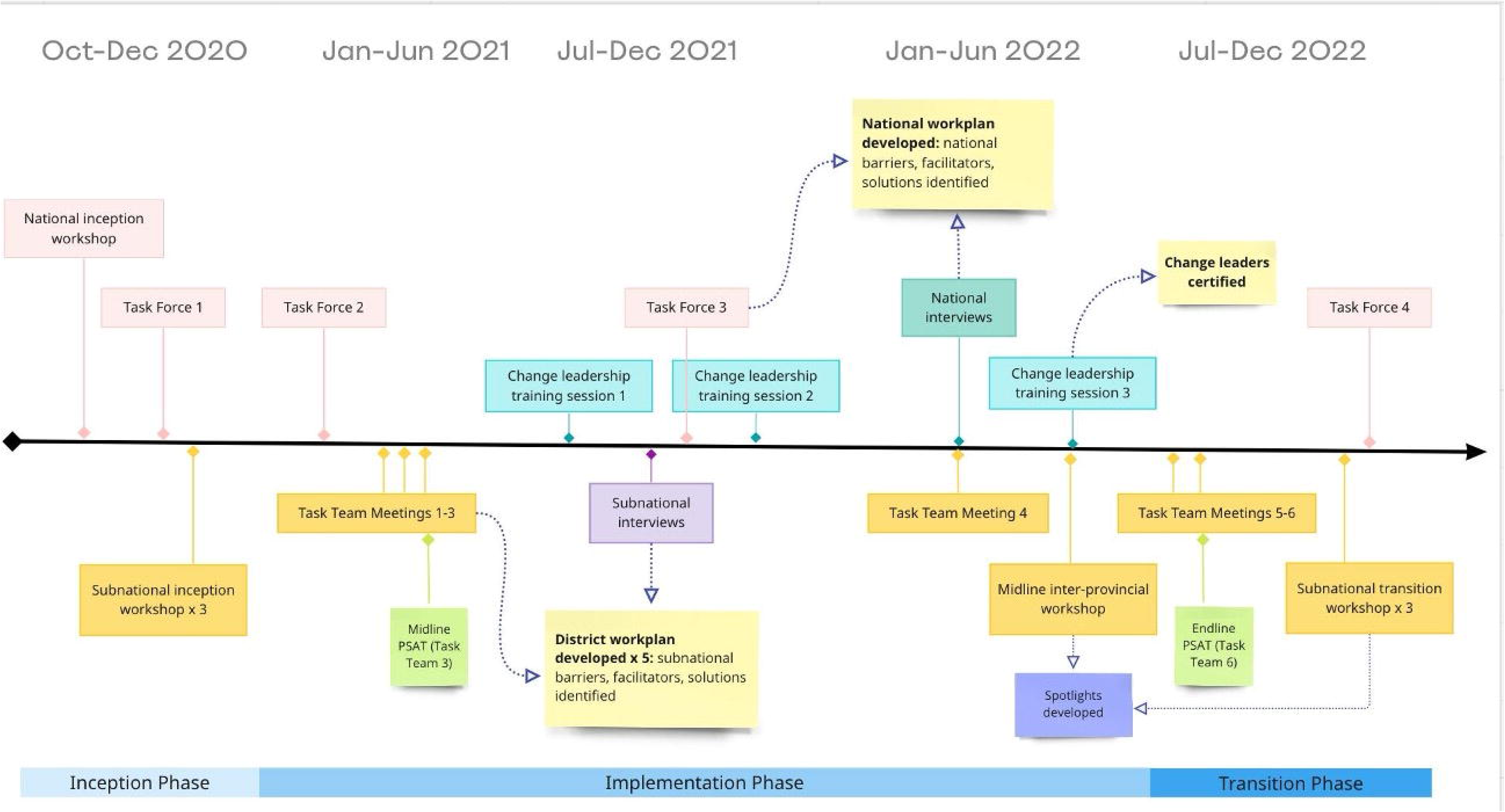
OPTIMISE project activities. Project activities took place at the national and subnational levels in Zimbabwe during a two-year period from December 2020-October 2022.

### Intervention areas

In close consultation with the MoHCC, the LEAD team selected three intervention provinces and five pilot districts, with the national VMMC Steering Committee’s approval (Table 1). The project partners considered the following factors in selecting participating provinces and districts: 1) representation of northern and southern regions; 2) stage of sustainability (scale-up or maintenance); 3) support from donor and implementing partners; and 4) accessibility by road (Fig 3).

**Table 1.** OPTIMISE project areas.

| Province | District | Funding Source | Implementing Partner |
| --- | --- | --- | --- |
| Matabeleland South | Matobo | PEPFAR/CDC<sup>†</sup> | ZAZIC Consortium (ZICHIRE<sup>‡</sup>) |
|  | Gwanda | PEPFAR/USAID | PSH<sup>§</sup> |
| Matabeleland North | Lupane | PEPFAR/CDC | ZAZIC Consortium (ZACH<sup>*</sup>) |
|  | Hwange | BMGF<sup>¶</sup> | PSH |
| Manicaland | Nyanga | Government of Zimbabwe/unfunded light-touch BMGF |  |

<sup>†</sup> Centers for Disease Control and Prevention
<sup>‡</sup> Zimbabwe Community Health Intervention Project
<sup>§</sup> Population Solutions for Health, Zimbabwe
<sup>\*</sup> Zimbabwe Association of Church-related Hospitals

**Fig 3:**
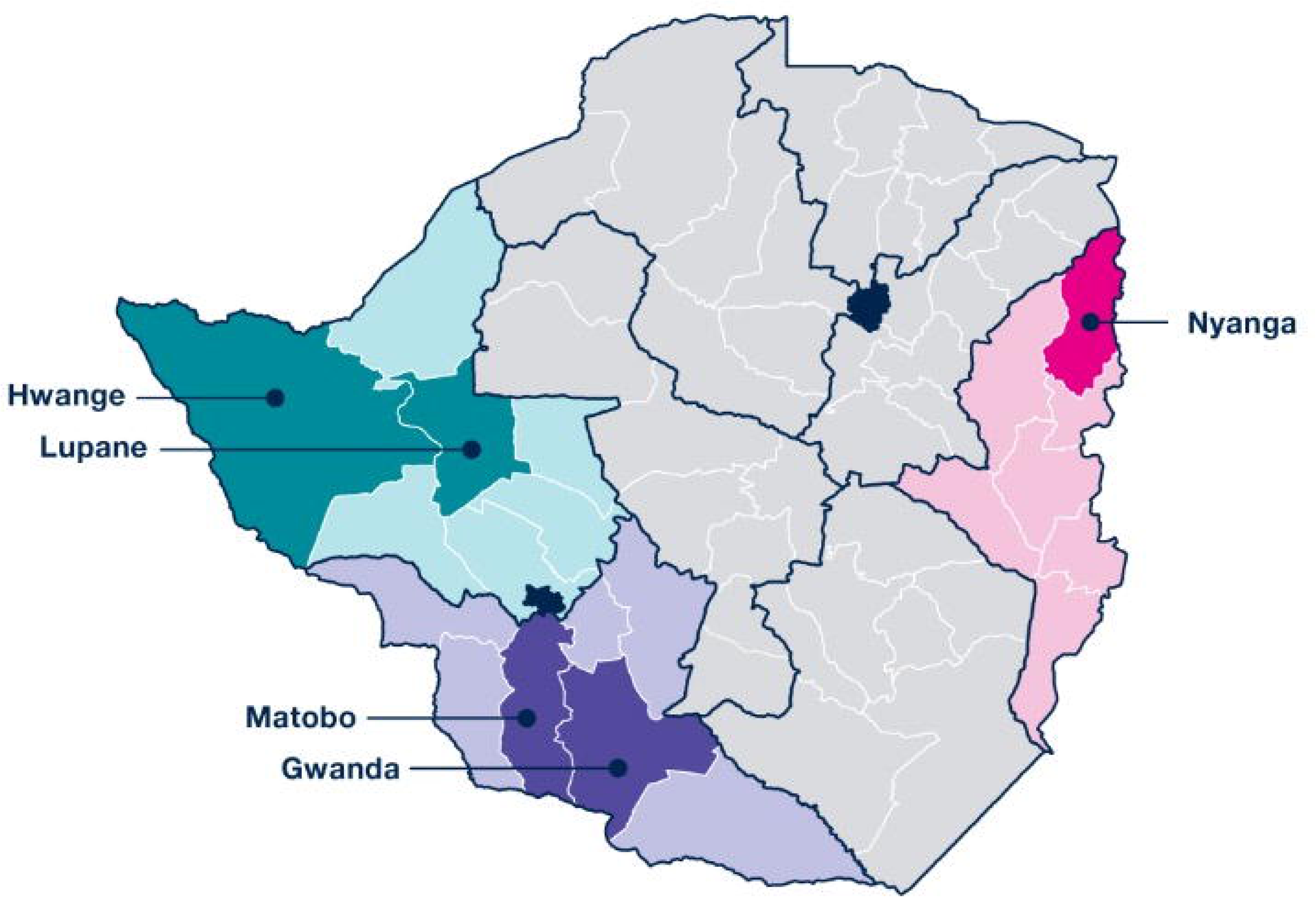
Map of OPTIMISE project districts. This map shows the three project provinces and their five corresponding districts (Hwange and Lupane in Matabeleland North; Gwanda and Matobo in Matabeleland South; and Nyanga in Manicaland.)

To illustrate the process of enhancing subnational capacity for leadership and management in the context of integrating a hybrid health program, we utilized a logic model, as depicted in Fig 4.

**Fig 4:**
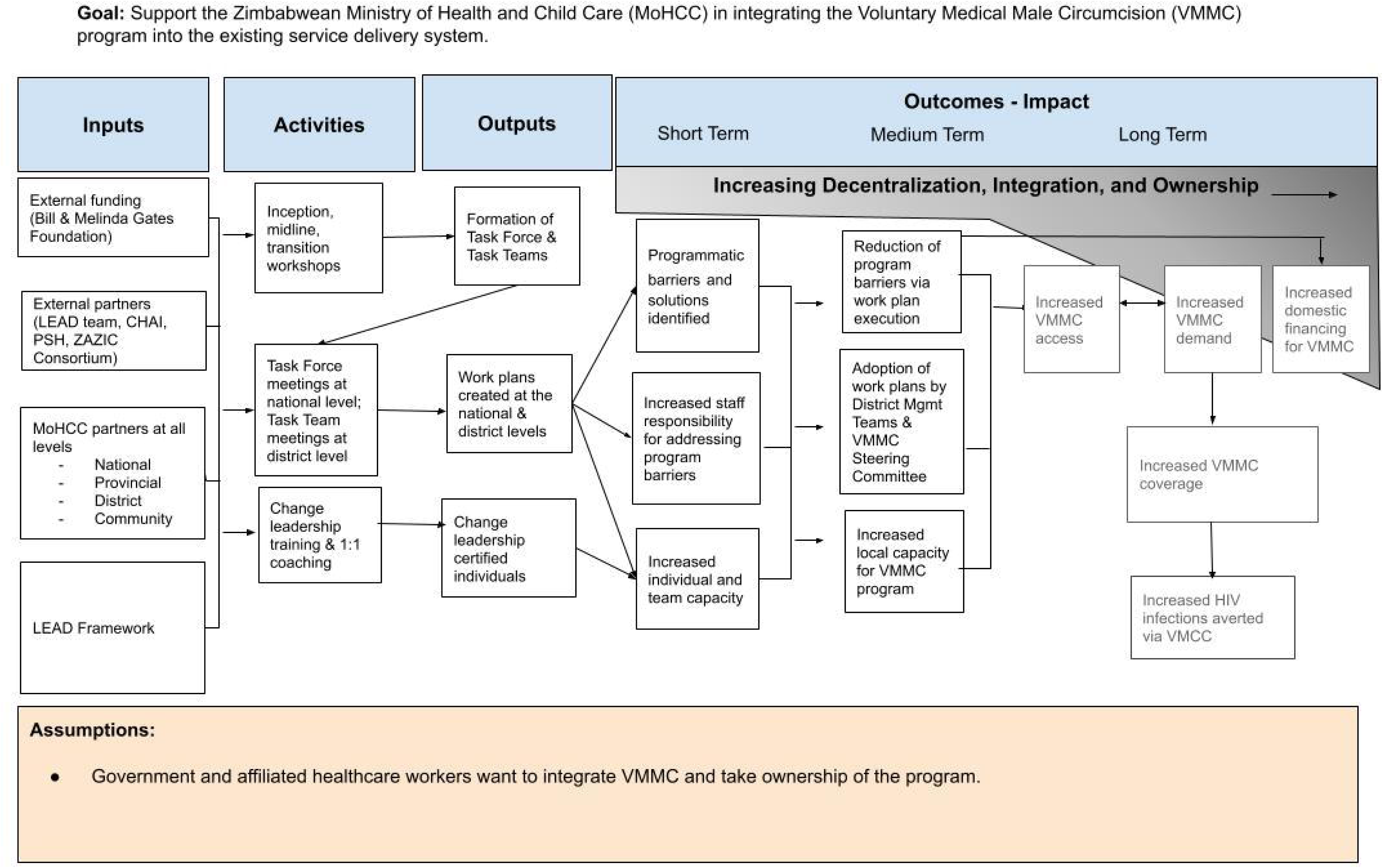
OPTIMISE project logic model.

### Outcomes

We will describe short-term and medium-term outcomes of the project. We present the first set of outcomes organized by the WHO Health Systems Building Blocks [34,35], a framework for describing a health system by six core components. These building blocks are critical components in the planning of sustainable VMMC services and provide a common language used across health programs [17].

### Evaluation data sources and analysis

We reviewed district work plans, assessing the progress made by the Task Teams in resolving the challenges to integration and sustainability by comparing baseline to endline quantitative and qualitative indicators. We determined that integration had been achieved when existing district management structures assumed responsibility for oversight of VMMC services, incorporating VMMC activities into their plans, budgets, and reviews. In addition to the district work plans, we collaborated on the identification of ‘Spotlights’. These were evidence-based change ideas for improving processes and strengthening the health system that could be replicated by others. Spotlights were generated by the five district Task Teams and served as evidence of how district teams adapted what they were doing to overcome the challenges of integration and sustainability.

We employed a Program Sustainability Assessment Tool (PSAT), a 40-item survey with 7-point Likert scale, administered anonymously at midline and endline to members of the Task Teams [36]. We collected midline survey data from a total of 54 respondents and from 59 respondents at endline. We aggregated PSAT results across all five districts. Since the surveys were administered anonymously, it was not possible to match scores to respondents. For each of the eight domains (e.g. organizational capacity), we calculated the mean midline and endline scores.

We collected evaluation data during project activities: inception, midline, and transition workshops, Task Force and Task Team meetings, and leadership development training sessions. We obtained ethical approval from the Medical Research Council of Zimbabwe (A2670), Research Council of Zimbabwe, and the University of California San Francisco Human Research Protection Program Institutional Review Board (20-39761). Ethical approval was given by all three entities.

## Results

All activities were implemented as planned from May 2020-October 2022, producing the following outputs: 1) formation of the national Task Force and district Task Teams, 2) national and district work plan development, and 3) a cohort of health professionals certified in change leadership (S2-S3 Tables).

### Integration and sustainability of VMMC

Our focus was on reporting medium-term outcomes that were achieved during a two-year time period. These outcomes involved actions by district Task Teams to reduce barriers and leverage opportunities for integration and furthering sustainability through work plan execution. demonstrating increased capacity of the VMMC program. We have organized these results by the WHO Health System Building Blocks and distilled them from the district level work plans (see Tables 4-7) (37). Additional results in the form of Spotlights are available at https://gatesopenresearch.org/documents/7-95 [37]. At the end of the project period, these work plans were institutionalized into existing structures by the District Health Executives and the national VMMC Steering Committee.

#### Leadership and governance results

Prior to our intervention, the implementing partner in each district oversaw the program without involvement by the District Health Executive (DHE), the governing body for all health programs within a district. At the end of our project, the DHE demonstrated increased responsibility for the VMMC program in all five districts. The inclusion of VMMC on regular meeting agendas indicated that these management structures had integrated VMMC into their consolidated district annual plans, making decisions about VMMC activities, budgets, and performance that they had previously left to their implementing partner (Table 4). In Hwange, VMMC was incorporated into a district platform used to coordinate and prioritize all development across sectors. This increased awareness and involvement, resulting in improved acceptance of VMMC at schools and communities, lead to the activation of 15 additional school health masters and improved community access by local leaders. In this same district, this change prompted increased sharing of budgets between programs and faster decision-making.

**Table 4.** District level leadership and governance results.

| Challenge | Solution | Indicator | Baseline | Endline | District |
| --- | --- | --- | --- | --- | --- |
| Limited participation by MoHCC representatives in the Rural District Development Committee (RDDC) | Use DHE funds from quarterly reimbursements to travel to meetings | VMMC on RDDC agenda (yes/no) | No | Yes: improved acceptance of VMMC at schools & communities resulted (increase of 15 school health masters activated for demand creation) | Hwange |
| DHE unaware of VMMC activities and budget | Ensure VMMC on DHE agenda & workplans | # of times VMMC on agenda/total # DHE meetings | 1/2 meetings (50%) | 3/3 meetings (100%): increased sharing of budgets between programs, faster decision-making | Hwange |
| Lack of integrated planning & review | Incorporate VMMC into meeting agendas | # of meetings where VMMC included/total # of meeting type | 0/5 (0%) | 5/5 (100%; Expanded Programme on Immunization, OI/ART, VIAC, RBF, DHE) | Gwanda |
|  |  |  | 0/2 (0%) | 2/2 (100%; DHE, District Management Team) | Lupane |
|  |  | # of meetings where VMMC included/quarter | 0/3 (0%) | 3/3 (100%) | Matobo |
| DHE not informed of VMMC activities | Add VMMC to DHE agenda, VMMC focal person, District Health Information Officer to share reports, regular review of VMMC data | # of meetings where VMMC is on agenda, data reviewed, & action taken | 0/quarter | 4/quarter | Nyanga |

#### Service delivery results

Evidence of integration of VMMC can be seen through improved accessibility and acceptability of services and subsequent increased performance (Table 5). Within Hwange district, the Task Team activated VMMC at a major town hospital that had not been offering VMMC and integrated it into its routine services, resulting in an increase in performance from 9 male circumcisions (MCs) per quarter at baseline during the height of the COVID pandemic to 101 MCs per quarter at endline (Table 3). In Gwanda, Lupane, Matobo, and Nyanga districts, the Task Teams empowered and engaged people to improve VMMC acceptability through village chief and religious leader dialogues, door to door campaigns, a music gala, a focus group with the target age group of males 15-29 years, and demand creation training for village health workers, school health masters, male champions, and adolescents. Nyanga also reoriented its service delivery model. To cut down on travel time and gas usage, VMMC teams were dropped off at locales closer to where clients lived, where they would camp and use local hospital vehicles for outreach travel. This change resulted in a doubling of outputs from 15 MCs/week at baseline to 30 MCs/week at endline.

**Table 5.** District level service delivery results.

| Challenge | Solution | Indicator | Baseline | Endline | District |
| --- | --- | --- | --- | --- | --- |
| Lack of VMMC services at two major hospitals in Hwange town | Engage management at hospitals to activate services | # of VMMCs performed by district facilities in town | 9 MCs per quarter | 101 MCs per quarter; remaining facility still needs to be activated | Hwange |
| Lack of program uptake | Hold quarterly meetings with key stakeholders | # of times key stakeholder meeting held/# of quarters | 0/4 (0%) | 4/4 (100%; 5 village chief community dialogues, door to door campaign, music gala, focus group with target age group) | Gwanda |
| Lack of community based demand creation | Identify stakeholders through mapping, conduct community trainings | # trained in demand creation<br># of times demand creation plans reviewed | 110/165 (67%) trained<br>0/52 (0%) plans reviewed | 160/165 (97%) trained<br>52/52 (100%) plans reviewed | Lupane |
| Insufficient demand creation due to COVID-19 | Conduct sensitization trainings | # individuals trained | 0 | 17 nurses<br>253 VHWs<br>22/24 District AIDS Action Committee (DAAC) members | Matobo |
| Limited MC outputs for fuel consumption, vehicle use | Review outputs for outreach visits, transport teams to camping spot, use local hospital vehicles for outreach travel | # of MCs per outreach visit/week | 15 MCs/week | 30 MCs/week | Nyanga |
| Religious barriers affecting VMMC uptake | Create demand platform through apostolic religious leaders | # of apostolic churches engaged | 0 churches | 10 churches |  |

#### Health workforce results

Expanding the health workforce that can perform VMMC is necessary to integrate it into routine services and imperative in Zimbabwe, where there are high levels of staff attrition. At the national level, the Task Force focused on the integration of VMMC in the pre-service nurse training curriculum, blended learning, and ways to expedite in-service certification. At the district level, all five Task Teams concentrated on training, certifying, and mentoring additional nurses (Table 6). Two districts (Gwanda and Nyanga) that were successful in certifying providers to work independently had access to a provincial trainer. In other districts that had to depend on verification by the limited number of national trainers, they dropped certification from their work plan or were not able to certify more than one circumciser (Hwange, Lupane, Matobo). In Lupane, the loss of 14 certified circumcisers meant that they had four fewer facilities that could offer VMMC. This district minimally increased its pool of trained circumcisers due to staff shortages and the reallocation of training funds to other activities. To compensate for these health workforce shortfalls and address the challenge, the Task Team created service delivery clusters for outreach teams. An innovative solution by the Matobo Task Team involved mobilizing funds from a local source to assist the DHE in providing supportive supervision to circumcisers. Nyanga merged their certification and conversion processes, resulting in the training of five more nurses and conversion of an additional four nurses. Formerly, the three step process of training, converting, and certifying health workers resulted in lag times of greater than 1 year.

**Table 6.** District Level Health Workforce Results.

| Challenge | Solution | Indicator | Baseline | Endline | District |
| --- | --- | --- | --- | --- | --- |
| Inadequate service delivery capacity due to resignation of 10 nurses | Train nurses from each facility | # of facilities with trained nurses/total facilities | 11/28 (39%) | 27/28 ( 96%) | Gwanda |
| Insufficient # of nurses certified | Certify nurses | # of nurse certified/# of nurses trained | 3/34 (10%) | 12/34 (35%) |  |
| Lack of supportive supervision | Mentor nurses | # of facilities where nurses mentored/total facilities | 0/28 (0%) | 9/28 (32%) |  |
| Staff attrition | Train health care workers | # of new HCWs certified/total HCWs certified | 0 new HCWs certified/25 HCWs certified (0%) | 1 HCWs certified/22 HCWs certified (5%)† | Hwange |
| Insufficient number of trained circumcisers due to attrition | Merged nurse conversion and certification processes | # of providers converted/total providers<br># of providers certified/total providers | Converted: 0/26 (0%)<br>Certified: 7/21 (33%)<br>Three step process of training, converting, and certifying health workers with lag times >1 year | Converted: 5/26 (7%)<br>Certified: 11/21 (52%)<br>Savings in fuel, time, per diem for trainers and greater client convenience | Nyanga |
| Insufficient number of RHCs with 2 trained circumcisers | Train & mentor HCWs | # of nurses trained/total nurses | 3/48 (6%) | 13/48 (27%); funds diverted from Training of Trainers, mentoring by partner | Matobo |
| Lack of funds for VMMC supportive supervision | Train DHE members on CQI tools to conduct quarterly visits, request funds from DAAC | DAAC contributions to supportive supervision | None | DAAC to contribute to transportation and lunch allowance for trainers |  |
| Attrition resulted in loss of 14 certified circumcisers (24→10), fewer sites with trained circumcisers (8/14→4/14) | Create service delivery clusters and outreach teams | # of clusters<br># of outreach teams with vehicle | 0<br>0 | 4 clusters<br>2 outreach teams with vehicles | Lupane |
| Insufficient number of trained circumcisers | Create duty roster, conduct refresher, conversion, basic trainings | # of duty rosters created<br># of HCWs trained (refresher, conversion, basic) | 0/12 rosters<br>refresher: 1/yr;<br>converted: 3/yr;<br>basic: 9/yr | 9/12 rosters created<br>refresher: 1/yr; converted: 6/yr; basic: 0/yr‡ |  |

†3 of the 4 healthcare workers who left the district were certified in VMMC.
‡Three rural health centers could not release their healthcare workers for training due to staff shortages. Furthermore, funds for basic training were reallocated.

#### Medicines, vaccines, and technologies results

Rather than using disposable surgical instruments that are costlier, wasteful, and affected by supply chain challenges, two districts focused on autoclaving to sterilize reusable instruments (Table 7). In Gwanda, the Task Team identified a local funding source to procure two additional autoclaves. In Hwange, the team formed a partnership with a private hospital, exchanging the use of a public hospital’s operating room and incinerator for access to the private hospital’s autoclave. This partnership resulted in an increase from sterilizing 15 packs/month in a quarter at baseline to 300 packs/month in a quarter at endline.

**Table 7.** District level medicines, vaccines, and technologies, strategic information, and financing results.

| Challenge | Solution | Indicator | Baseline | Endline | District |
| --- | --- | --- | --- | --- | --- |
| Availability of autoclaves at all facilities | Identify source of funding and procure autoclaves | # of facilities with autoclaves/total facilities | 26/28 (93%) | 28/28 (100%) | Gwanda |
| Insufficient autoclaving capacity | Formed partnership with private hospital in exchange for incinerator and operating theater access | # of VMMC packs available/quarter | 15 packs per month/quarter | 300 packs per month/quarter | Hwange |
| Late, incorrect, incomplete facility data | Ensure all facilities have forms, conduct quarterly on-site data verification, mentoring, collected data at the same time as lab specimens | % of sites visited/total sites<br>% of sites reporting on time | 3% (1 site visit/32 sites)<br>60% sites (19/32) reporting on time | 91% (29 site visits/32 sites), reduced time needed to follow-up about incomplete data, improved data quality<br>100% sites (32/32) reporting on time | Nyanga |
| Incomplete facility data | Discuss data quality gaps with all sites, provide mentoring | % completeness of monthly return forms | 54% complete/4 sites | 100% complete/4 sites | Gwanda |
| Power interruptions affecting data quality | Identify funds to procure fuel for generators | GOZ funds mobilized | \$0 | US\$1,120 | Nyanga |
| Inoperable vehicles, lack of fuel for VMMC | Identify funds for vehicle servicing and fuel, create servicing schedule | Funds mobilized<br>Schedule developed | Servicing: \$0<br>Fuel: \$0<br>Schedule: no | Servicing: US\$4,000/year<br>Fuel: US\$1,792/year<br>Schedule: yes | Nyanga |
| Insufficient funds for VMMC activities | Allocate GOZ funding towards VMMC | Funds for outreach, fuel, mentoring | 0 | 10% (funds for fuel were diverted towards emergencies) | Matobo |

#### Strategic information results

The Nyanga and Gwanda Task Teams focused on VMMC data quality at facility level. To address late, incorrect, incomplete data, the Nyanga team conducted quarterly on-site data verification and ensured forms were supplied to all health facilities and that monthly return forms were transported to the district with routine delivery of lab specimens. Their supportive supervision visits increased from conducting 3% of total site visits/year at baseline to 91% of total site visits/year at endline (Table 7). Timeliness of monthly return form submissions increased from 60% of facilities (19/32 sites) to 100% of facilities (32/32 sites). The Gwanda management team went from conducting separate VMMC data verification visits to initially covering all program areas and then later integrating VMMC into more comprehensive quarterly results-based financing supervision visits. This team also focused their efforts on four facilities with incomplete data to address accuracy and reduce variance between primary data sources and monthly return form reports. By discussing the data quality gaps, mentors at these facilities improved completeness of forms from 54% to 100% at all four sites.

#### Financing results

Vehicles and fuel to transport VMMC outreach teams is challenge common across districts, due to the use vehicles for other health program activities. Some districts have donor-funded vehicles. In the absence of such a vehicle, the Nyanga Task Team identified local funds for vehicle servicing and fuel and created a vehicle servicing schedule for the fleet of vehicles it uses for all health programs, demonstrating integration of VMMC. It went from having no funds for these expenses and no schedule at baseline to having a budget of US$4,000/year for vehicle servicing and US$1,792/year for fuel and a regular servicing schedule at endline.

Additionally, Nyanga district mobilized local funds to prevent power interruptions affecting data quality. They identified funds to procure fuel for generators, with no budget at baseline to US$1,120 as a dedicated budget line item for generator fuel. In Matobo, the district team allocated Government of Zimbabwe (GOZ) funding towards VMMC activities. At baseline none of the DHE budget funded VMMC, while at endline, 10% of the total DHE budget was used for VMMC outreach and mentoring. The share of the budget for VMMC would have been larger if the DHE had not diverted funds for fuel elsewhere (Table 7). In both districts, the DHEs mobilized local funding towards VMMC, showing progress towards sustainable financing.

### Program sustainability survey results

Increased capacity to identify programmatic barriers, opportunities, and solutions was further supported by the results of a program sustainability assessment. An additional output of localized definitions of integration and sustainability enabled the district teams to build consensus on what they were trying to achieve (S4 Table). We examined progress towards program sustainability using a standardized Program Sustainability Assessment Tool (Fig 5) to supplement the work plan results. By aggregating the results across all five districts, calculating the means for each domain, and performing Mann-Whitney tests, we found a significant difference between midline and endline measurements for three of the eight domains: communications (p=0.05), program adaptation (p=0.05), and organizational capacity (p=0.02). The most substantial increases in absolute values were in the domains of communications (+0.05), program adaptation (+0.04), organizational capacity (+0.04), and partnerships (+0.04). The only decrease in absolute values between midline and endline results was in funding stability (−0.04). Reasons for the decline in financial sustainability varied across districts, including reduction of funding levels, limited flexibility to shift funds to emerging priorities, a flat rate per circumcision, regardless of distance travelled or fuel consumed, reductions in rates for mobilizers, and delays in financial disbursements. An example of increased organizational capacity that extended beyond the HIV program is described in S5.

**Fig 5.**
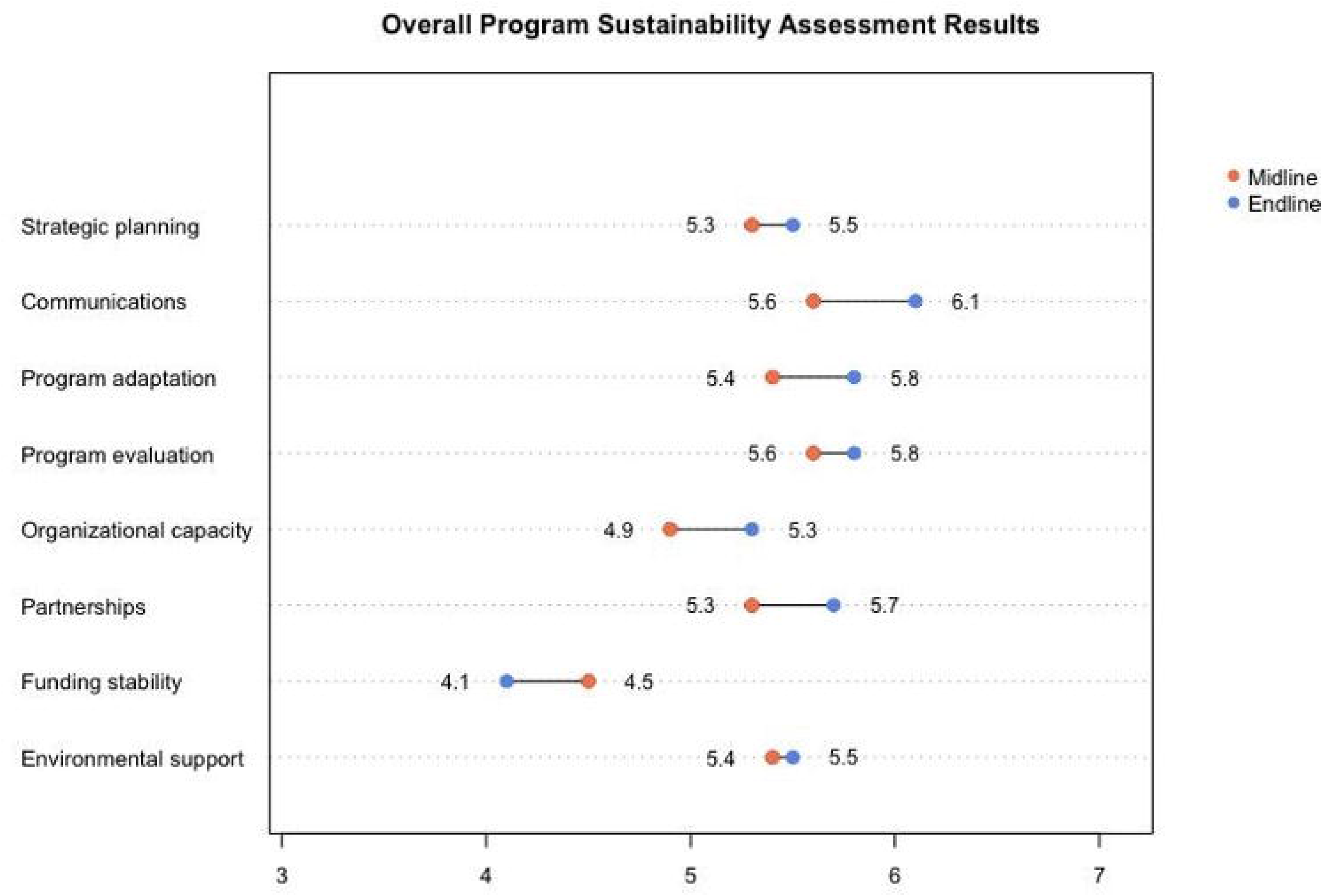
Overall program sustainability assessment results. Substantial increases in absolute values between midline and endline mean measurements were in communications, program adaptation, organizational capacity, and partnerships. The only decrease was in funding stability.

## Discussion

We set out to integrate the VMMC program into routine services while also furthering sustainability in five pilot districts. Through application of the LEAD Framework, we facilitated transition readiness and a change in mindsets within the five pilot districts that VMMC should be a locally-owned and managed intervention. LEAD also strengthened individual and team capacity. District teams assumed the planning, oversight, and budgeting of VMMC, furthering the integration of the VMMC program and strengthening the health system. They also raised financing for themselves from local funds. District team actions included greater engagement of multisectoral stakeholders, better use of existing resources, and changing operating models. The approach also introduced a more effective way of articulating and resolving granular VMMC challenges at the district level with management and oversight at provincial and national levels. LEAD resulted in improved system communications and management for VMMC services.

The VMMC program was integrated with other health services to the extent that existing district management structures took responsibility for oversight of VMMC services, incorporating VMMC activities into their plans, budgets, and reviews. In one district, further decentralization occurred, with the delivery of VMMC through two additional health facilities that provide routine health services. However, across the five districts, there was an overall reduction in the number of static health facilities offering VMMC. At the same time, service delivery at all general health facilities is not necessarily desirable nor attainable with such high levels of staff attrition. Despite this drop in the total number of static health facilities, the total number of male circumcisions performed increased over this period by clustering service delivery areas.

Lessons from Kenya, which began its process earlier than Zimbabwe, underscore that a purely static facility model is insufficient for achieving integration and sustainability. A 2021 study demonstrated that a mixed model—where circumcisers offer both static and mobile (outreach) services—proved more efficient and effective compared to static models alone [38]. Both this study and a separate case study from Kenya highlighted inadequate domestic financing as a significant barrier to the country’s progress towards sustainability [39]. Moreover, achieving integration and sustainability requires time, and focusing exclusively on circumcising adolescents and men is unlikely to suffice. The case study also identified the lengthy integration of services into public health facilities and limited support for infant male circumcision as additional challenges.

The OPTIMISE project was successful in facilitating improvements across all the WHO health system building blocks, suggesting that the intervention strengthened the overall health system and supporting the idea of taking a diagonal approach [26,27]. In all five districts, accountability for the VMMC program by District Health Executives increased. Due to staff attrition, the continual need to train the in-service health workforce did not always yield a sizable increase in circumcisers. However, a reorientation of services demonstrated program adaptation. These actions also reduced vehicle and fuel use and staff travel time, contributing to an overall increased volume of MCs performed. Creation of a duty roster was another innovation that could be used for other health services to create greater equity and staff satisfaction. More attention should be focused on addressing staff burnout and attrition, examining reasons for dissatisfaction and departures, and considering incentives to mitigate these problems. These factors affect the entire health system, and Zimbabwe is just one of many low- and middle-income countries where health worker migration has serious adverse effects [40]. Furthermore, considering the interest to sustain the VMMC program through 2030 and insufficient domestic resources, donors should continue to invest in this important HIV prevention strategy in Zimbabwe.

While a higher education institution was involved in adapting the LEAD Framework for this project, scaling of the approach can be done by trained local facilitators. After the initial pilot stage of this project, these local facilitators have expanded the approach to additional districts, at an approximate cost of US$30,000 per district. The cohort of trained change leaders within Zimbabwe can support these expansion efforts.

### Limitations

There were several limitations to our project. First, we were not able to measure performance across districts using a common set of metrics because we gave the district teams the agency to develop their own indicators, resulting in variation across the five districts. While we did gather data on trained circumcisers, facilities offering VMMC, and overall VMMC performance, this was either incomplete or not consistently uniform, which prevented comparison across most parameters. Second, the standardized program sustainability survey was not administered at baseline. Therefore, we only have comparisons of midline and endline data. Third, one district could not improve drug stock outs, and our project could not influence this, due to shortages at the national level. Fourth, we did not determine whether focused efforts on the VMMC program influenced the performance of other health programs. Although we suggest that the LEAD Framework contributed to the results, we cannot be certain about attribution without doing an impact evaluation. We were also not able to isolate the impact on project outcomes of COVID-19 disruptions in VMMC service delivery and subsequent resumption of services. Finally, we conducted an internal evaluation, where the funder supported both this project and the evaluation.

## Conclusion

Our approach is worth consideration by other vertical health programs and low- and middle-income countries that have a goal of integrating and sustaining health services while strengthening their health systems using a bottom-up model that prioritizes localization. Readers interested in applying a similar approach can refer to the LEAD Framework User Guide (https://shrinkingthemalariamap.org/sites/default/files/tools/lead-user-guide-final-updated-12-7-2021.pdf).

LEAD should be scaled to other districts within Zimbabwe and considered by other donors for strengthening leadership and governance and maximizing health program effectiveness and efficiencies. However, further work is needed to identify funding sources for expert facilitators within mainstream health services and formalizing the levels of expertise required for program facilitation, like that used by LEAN/Six Sigma. Shifting ownership of the VMMC program in Zimbabwe to local stakeholders will require a change in who provides and controls the funding for VMMC. Beginning in 2018, PEPFAR set explicit targets for shifting financial control from external to local entities [41]. This needs to be coupled with greater domestic resource mobilization for HIV, which will contribute to sustaining the program until 2030, especially given the flatlining of funding for HIV prevention. Donors should continue to support the investment they made in strengthening governance of the health system and leverage the momentum of empowering and further capacitating government stakeholders to lead the program. That way, the VMMC program can continue to serve as an important entry point for males who might not otherwise engage with the health system while also contributing to ending the HIV epidemic.

## Data Availability

All data produced in the present study are available upon reasonable request to the authors.

## Acknowledgments

The authors wish to thank Tatenda Ruwuyu, Katie Joyce, Priscilla Mataure, and Greyling Viljoen, Task Force Leader Dr. Simon Nyadundu, Provincial Medical Directors for Manicaland, Dr. Mukuzunga, Matabeleland North, Dr. Kuretu, and Matabeleland South, Dr. Chikodzore, PMCHO for Manicaland Dr. Nyafesa, and the Task Teams in Gwanda, Hwange, Matobo, Lupane, and Nyanga.

