## Supplemental Table 1 for "Sustainable integration of a vertical voluntary medical male circumcision program into routine health services in Zimbabwe: A mixed methods evaluation of a participatory change intervention"

### **S1 Table: LEAD Framework techniques and tools**

| **Technique** | **Description / examples of tools** |
| --- | --- |
| Participatory Action Research (PAR) | A specific set of interventions designed to enhance healthcare service delivery and organizational effectiveness. Where organizing is seen as a process that requires continuous and numerous activities, PAR enhances capacity for crucial aspects of human relations and activities – typically the softer aspects that rely on such qualities as listening, respect, reflection, and adapting to political realities [35]. PAR interventions often include structured techniques designed to evince insight from frontline and community-based stakeholders; to engage line managers in responding to these insights; and to embed accountability for change at all levels of the system. Examples are a) peer-led problem solving, b) attentive listening, c) process mapping, and d) assessment of inter-group dynamics. |
| System in the Room | A methodology used to ensure that a representative set of stakeholders who need to be engaged in transition of the VMMC program are present at key workshop events. Originating in the fields of psychosocial studies and organizational dynamics, it entails replicating the program/service delivery system as fully as possible in a shared workshop setting (e.g., a conference room) [44]. Representation includes not only the healthcare professionals at district level who are responsible for service provision, but also more senior staff and resource holders from provincial and ministry levels, community decision-makers, and intended beneficiaries. Full representation enables sharing of perspectives and challenges from across the system and helps inform collaborative generation of challenges, synchronized solutions and collective support for those whose role it is to implement frontline solutions involving changes in processes and procedures. Ensuring those with the seniority to authorize and resource changes is critically important to the process, from national to community levels, so that varying perspectives and experiences were considered in grappling with the barriers to integration and sustainability of the program. The methodology also enables real-time appraisal of program delivery dynamics and cross-system generation of solutions to the challenges identified by stakeholders. |
| Friendly Consulting | A structured way of generating fresh insight into healthcare problems in small groups. It is a peer-led process, typically involving five or six individuals who take on designated ‘client’ (challenge presenter) and ‘consultant’ (feedback giver) roles. The consulting process for each challenge takes forty-five minutes after which roles change. Each person in the group has an opportunity to present and get feedback on a challenge they bring to the group. This technique was used repeatedly to facilitate peer-led problem solving in small groups on VMMC challenges. |
| Facts and Feelings | A structured small group (three person) exercise that encourages participants to listen actively and facilitates interpersonal communication in intervention workshops and meetings. |
| Root cause analysis | 5 whys, fishbone diagrams, and process mapping |
| Solution generation | Brainstorming and reverse brainstorming |
| Prioritization | Pareto analysis and effort versus impact matrices |
| Professional Practice in Change Leadership (PPCL) sessions | The module consisted of experiential/work-based learning that ran concurrently with project activities and was supported by a series of three face-to-face or blended learning workshops spaced evenly across the academic year. Students were tutored by LEAD experts in the application of the organization development facilitation methods and quality improvement techniques that comprise the LEAD Framework. In addition, students completed a series of reading assignments and written work that culminated in two summative assessments: submission of a personal learning portfolio and completion of an independent project in which students reported on up-scaling or out-scaling work they had completed in their own specialist areas.  Content covered during sessions: 1) Introduction to organization development (OD) and process improvement in healthcare programs, implementing OD processes, participant roles and responsibilities; 2) Presentations: implications of leadership learning and its application to a change project. Reading assignment review and application of principles. Theory and practice: Advanced facilitation processes, skills and techniques. Identifying opportunities to extend the OD approach into the workplace; 3) Final project presentations: review of practice, outcome and output measures, role and authority of facilitators. Personal and group feedback. |
