## Supplemental Tables 2, 3, 4 for "Sustainable integration of a vertical voluntary medical male circumcision program into routine health services in Zimbabwe: A mixed methods evaluation of a participatory change intervention"

**S2 Table. District Task Team composition**

| **Type** | **Organization/ Program** | **Roles** |
| --- | --- | --- |
| Community | Churches, local government | Pastor, Village Chief, Councillor |
| Facility | MoHCC | Registered General Nurse, Pharmacist |
| District | MoHCC, Ministry of Primary & Secondary Education, local government | District Medical Officer, District Nursing Officer, District Health Promotion Officer, District Health Services Administration Officer, District AIDS Coordinator, District Health Information Officer, District Schools Inspector, District Development Coordinator |
| Provincial | MoHCC | Provincial Medical Director, Provincial Nursing Officer, Provincial Maternal and Child Health Officer, Provincial VMMC Officer, Provincial Health Promotion Officer |
| National |  | AIDS and TB Program Officer, TB Officer, Data & Research Officer, HIV Prevention Coordinator, Program Officer, Monitoring & Evaluation Officer |
| Implementing/ technical partners | CHAI, PSH, ZIM-TTECH, ZACH, ZiCHIRe | Management Development Specialist, Analyst, Deputy Director, HIV Prevention Manager, VMMC Demand Creation Manager |

**S3. National Task Force composition**

| **Type** | **Organization/ Program** | **Roles** |
| --- | --- | --- |
| National | MoHCC, National AIDS Council | AIDS and TB Program Officer, TB Officer, Data & Research Officer, HIV Prevention Coordinator, Training Officer |
| Provincial | MoHCC | Provincial Medical Director, Provincial Epidemiology and Disease Control Officer, Provincial Maternal and Child Health Officer |
| Implementing & technical partners, donors | PSH, ZIM-TTECH, WHO, CHAI, BMGF, PEPFAR (CDC, USAID), Chemonics | Director, Deputy Program Director, Program Manager, National Professional Officer, Senior Analyst, Country Liaison, HIV Prevention Manager |

**S4 Table: Examples of localized integration and sustainability definitions**

| Term | Definition |
| --- | --- |
| Sustainability | The continuity of quality VMMC services as part of a comprehensive HIV prevention strategy and recognition of VMMC as an essential service, which must be available in all health institutions at all times with full ownership by all stakeholders. |
| Integration | The recognition of VMMC as part of the package of essential core health services at primary and secondary levels of care which is offered to all eligible clients, together with other health services, at an affordable cost, efficiently utilizing available resources. |
