## Supplemental Table 5 for "Sustainable integration of a vertical voluntary medical male circumcision program into routine health services in Zimbabwe: A mixed methods evaluation of a participatory change intervention"

**S5 File: Example of increased organizational capacity beyond HIV program**

Despite having an OB/GYN specialist in the district, Hwange District reported 9 maternal deaths in 2021, the highest maternal mortality rate in Matabeleland North province. The District Medical Officer (DMO) was determined to do something to improve this situation. She knew how demoralizing and traumatic it was for the Victoria Falls Hospital staff every time a mother died while giving birth. As the head of the District Health Executive (DHE), the DMO worked with her team to employ root cause analysis and then determine a plan of action. Root cause analysis is a set of tools she had learned through the OPTIMISE project. As a team, the DHE identified two factors contributing to the problem: 1) the lack of a blood bank and 2) a shortage of labor and delivery drugs such as misoprostol. Without a blood bank, Victoria Falls Hospital would send a vehicle 100 kilometers away to retrieve blood bags when they had a patient with postpartum hemorrhage. In the 3 hours they had to wait for blood, the hospital staff was losing the mothers who had just delivered their babies. To address the situation, the DHE engaged the national blood service, appealing the decision to not have a blood bank at the hospital. At the same time, the DMO ensured that the hospital’s blood storage refrigerator was fixed. She also required that the hospital maintain an adequate supply of misoprostol. With these changes in place, the district has reported 3 maternal deaths in 2022.
