## Supplemental Table 6 for "Sustainable integration of a vertical voluntary medical male circumcision program into routine health services in Zimbabwe: A mixed methods evaluation of a participatory change intervention"

**S6 Table: List of abbreviations**

| AIDS | Acquired immunodeficiency syndrome |
| --- | --- |
| ART | Antiretroviral therapy |
| BMGF | Bill & Melinda Gates Foundation |
| CDC | Centers for Disease Control and Prevention |
| CHAI | Clinton Health Access Initiative |
| DAAC | District AIDS Action Committee |
| DAH | Donor assistance for health |
| DHE | District Health Executive |
| DHIO | District Health Information Officer |
| DMT | District Management Team |
| EPI | Expanded Programme on Immunization |
| GOZ | Government of Zimbabwe |
| HCW | Healthcare worker |
| HIV | Human immunodeficiency virus |
| HPV | Human papillomavirus |
| IP | Implementing partner |
| LEAD | Leadership and Engagement for Improved Accountability and Delivery of |
| MC | Male circumcision(s) |
| MoHCC | Ministry of Health and Child Care |
| OI | Opportunistic infection |
| PEPFAR | U.S. President’s Emergency Fund for AIDS Relief |
| PMCHO | Provincial Maternal and Child Health Officer |
| PMD | Provincial Medical Director |
| PPCL | Professional Practice in Change Leadership |
| PSAT | Program Sustainability Assessment Tool |
| PSH | Population Solutions for Health |
| PSI | Population Services International |
| QI | Quality improvement |
| RBF | Results based financing |
| RDDC | Rural District Development Committee |
| RHC | Rural health centre |
| STIP | Sustainability Transition Implementation Plan |
| STI | Sexually transmitted infection |
| TB | Tuberculosis |
| UCSF | University of California San Francisco |
| VIAC | Visual inspection with acetic acid and cervicography |
| VMMC | Voluntary Medical Male Circumcision |
| VTAD | VMMC Transition Assessment Dashboard |
| WHO | World Health Organization |
| ZAZIC | ZACH, ZICHIRE, and Zim-TTECH consortium |
| ZACH | Zimbabwe Association of Church-related Hospitals |
| ZiCHIRE | Zimbabwe Community Health Intervention Project |
| Zim-TTECH | Zimbabwe Technical Assistance, Training, and Education Center for Health |
